# How do growth and nutrition explain social inequalities in lung function in children with cystic fibrosis? A longitudinal mediation analysis using interventional disparity effects with time-varying mediators and intermediate confounders

**DOI:** 10.1101/2022.01.11.22268909

**Authors:** Daniela K. Schlüter, Ruth H. Keogh, Rhian M. Daniel, Schadrac C. Agbla, David Taylor-Robinson

## Abstract

**Background:** Deprivation is associated with poorer growth, worse lung function and shorter life expectancy in children with cystic fibrosis (CF). While early growth is associated with lung function when first measured at around age 6, it is unclear whether improving early growth in the most disadvantaged children would reduce inequalities in lung function.

**Methods:** We used data from children born 2000-2010 and followed up to 2016 in the UK CF Registry. To estimate the association between deprivation and lung function at around age six, and the causal contribution of early weight trajectories, we extended the mediation analysis approach based on interventional disparity effects to the setting of a longitudinally measured mediator. We adjusted for baseline confounding by sex, birthyear and genotype and accounted for time-varying intermediate confounding by lung infection.

**Results:** 853 children were included in the study, including 165 and 172 children from the least and most deprived population quintiles, respectively. The average difference in lung function between the least and most deprived quintile of children, was 4.51 percent of predicted forced expiratory volume in one second (95% CI: 1.08 - 7.93). We estimated this would be reduced to 3.97 percentage points (95% CI: 0.57 – 7.38) if early weight trajectories in the most deprived children were shifted to the distribution observed in the least disadvantaged children.

**Conclusion:** Socio-economic conditions are strongly associated with lung function for children with CF which we estimated would only be marginally reduced if early weight trajectories could be improved for the most disadvantaged children.

## Introduction

Cystic fibrosis (CF) is a serious inherited condition. 1 in 2,500 babies born in the UK have CF, with over 10,000 people living with CF in the UK today. CF is caused by variants of the CF transmembrane conductance regulator (CFTR) gene, resulting in the build-up of thick, sticky mucus in various organs, in particular those of the respiratory and digestive systems. Most people with CF die prematurely from their disease through respiratory failure.

Growth and nutrition in early life are associated with lung function at age six for children with CF and interventions aimed at improving growth in early life may improve later pulmonary function^1^. The respiratory system develops from shortly after conception until adolescence and matures most rapidly in size and intricacy in the first three years. Therefore, adequate nutrition during this phase is deemed essential. In children with CF diagnosed within 6 months after birth and who were malnourished at diagnosis, catch-up growth by age two is associated with improved lung function at age six, while maintenance of growth from ages two to six has no strong additional effect on pulmonary function^2^.

Children with CF from socio-economically disadvantaged areas in the UK have worse outcomes than those from more affluent areas^3–5^. Socio-economic inequalities in weight and nutritional status have been reported from infancy and inequalities in lung function are evident as soon as this can be routinely measured at around age six^3^. Although there is some evidence from other countries that inequalities may widen with age^6,7^, no evidence has emerged that this is the case in the UK^8^. Therefore, understanding the pathways that lead to inequalities in lung function at age six is crucial to identifying potential targets for interventions that could reduce inequalities in CF across the lifecourse.

Achieving and maintaining healthy growth and nutrition is difficult for many people with CF due to fat malabsorption, increased energy requirements, reduced oral energy intake and psychosocial factors^9^. Thus dietary management has been found to be a difficult task for children with CF and their parents^10^, further compounded by socio-economic adversity. Studies in the USA have shown that parental CF-specific nutritional knowledge is a strong predictor of nutritional outcomes^11^ and also strongly associated with parental income and level of education^12^. In addition, there has recently been an increased awareness of high levels of food insecurity in people with CF in the USA^12,13^. Patterns are likely similar in the UK although there are currently no data on the number of children with CF that live with food insecurity.

In this study we examined how growth and nutrition in the first six years of life can explain inequalities in lung function (when first measured around age six) between groups with different levels of deprivation in the UK CF population. More specifically, we were interested in estimating the association between deprivation at birth and lung function at around age six, and the extent to which this could be reduced if we were to intervene to improve the weight trajectories in the most deprived children such that they have the same distribution as those in the least deprived children.

To estimate the effects of interest, we build on recent developments in the causal inference literature that have led to the definition of “interventional disparity effects”^14^ and extend this approach to the mediation setting with a longitudinally measured mediator.

## Methods

### Data Source

We used UK CF Registry data, which collects data at approximately annual clinic visits when a patient is clinically stable. Records date back to the 1990s and are estimated to capture 99% of the current UK CF population^15^. Our data included follow-up to the end of 2016. Annual review visits include evaluation of clinical and nutritional status, lung function, and microbiology of respiratory tract secretions since the last annual review. Demographic data, including socio-economic status, are also recorded.

### Setting and Participants

We included children who were born after 2000, diagnosed by newborn screening, had at least one lung function measure between ages ≥6 and <9 years and at least one weight measurement between birth and <7 years of age, i.e. before or at the same time as the lung function measure. We excluded individuals who did not have complete data on sex, genotype, year of birth and socio-economic conditions (SECs).

### Outcome, exposure, mediator, and confounders

Our outcome of interest was the first available lung function measurement given by percent of predicted forced expiratory volume in one second (%FEV1) based on GLI reference equations, which adjust for sex, age, height and ethnicity^16^(continuous variable).

Our exposure of interest was SECs at birth. Measures of individual SECs are not available in the UK CF Registry. Therefore, we used the Index of Multiple Deprivation (IMD), a small area deprivation measure, based on first home-address postcode recorded in the registry, as proxy for SECs at birth. The IMD is a measure of relative deprivation which combines information on income, employment, health, education, access to services, crime/safety and environment/housing^17–20^. The domains vary between England, Wales, Scotland and Northern Ireland. In order to obtain a comparable measure of deprivation across countries, we calculated country-specific IMD z-scores^5^. All children were included in the study but our main interest was the difference in outcome between children in the highest and lowest quintile of deprivation (see Supplementary Material for details). We therefore converted z-scores to quintiles.

Our mediator of interest was early weight measured at annual review visits from birth up to but excluding age 7. We used weight-for-age z-scores based on the UK WHO reference^21^ as a continuous variable.

Confounders were identified based on our assumed causal relationships between SECs, weight, lung function and their common causes (Figure 1). We included sex (male/female), birthyear (continuous) and genotype (categorical: number of F508del alleles). Respiratory infection may be a time-varying confounder of the mediator-outcome relationship affected by the exposure^3^. We only considered infection by *Pseudomonas aeruginosa*, the most common cause of infection in the UK CF population under study^22^. At any annual review visit, it is recorded whether an infection with *P. aeruginosa* had been present since the last annual review visit.

**Figure 1:**
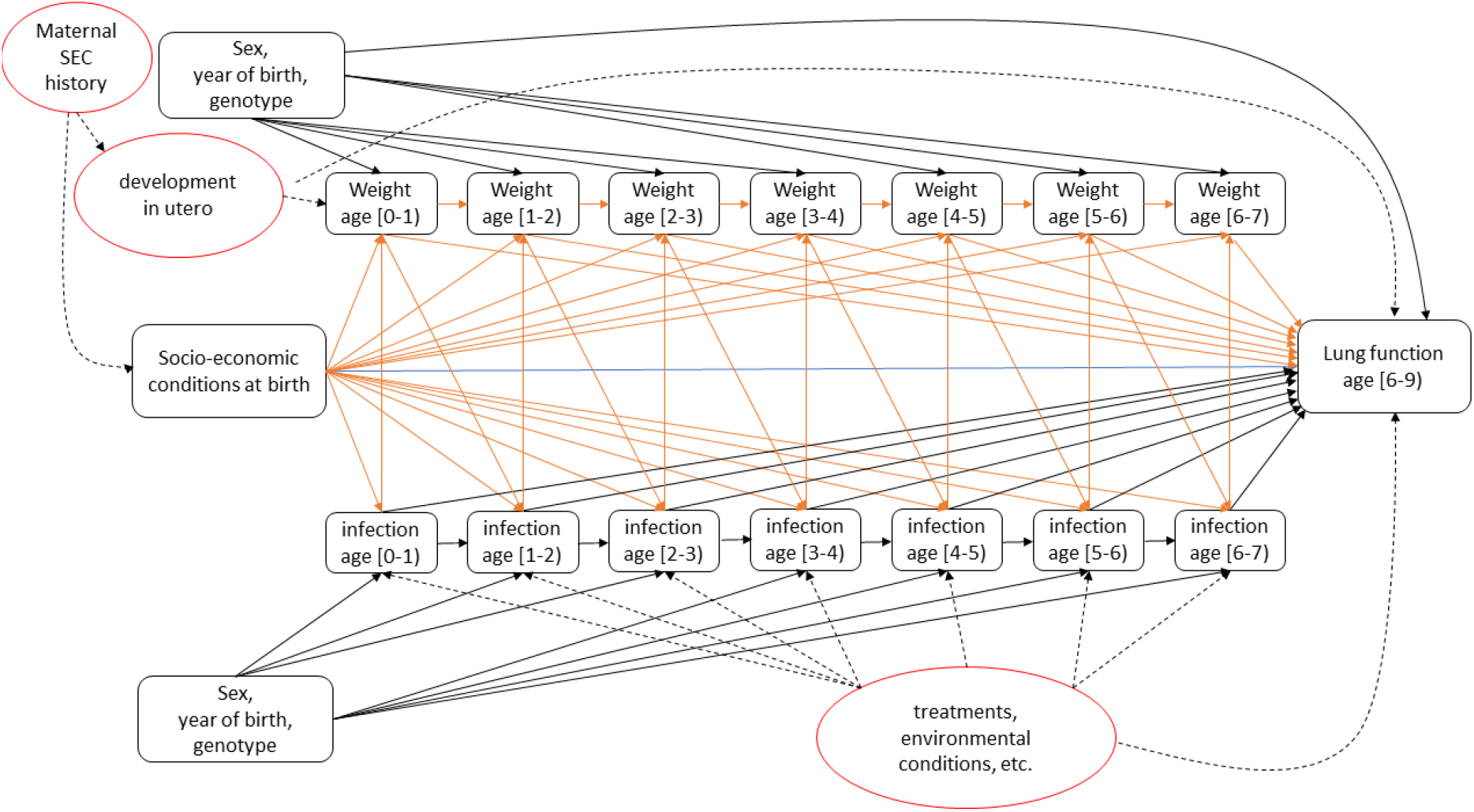
Assumed causal relationships between socio-economic conditions at birth, early weight, first lung function measurement and their common causes including baseline and time-varying confounders of the mediator-outcome relationship that are also affected by exposure, and latent variables, which are assumed present but not observed (red ellipses).

### Statistical methods

#### Rationale and effects of interest

We were interested in estimating the association between SECs at birth (high versus low) and lung function at around age six and the extent to which it can be explained by early weight trajectories. Figure 1 illustrates the assumed causal relationships between SECs, weight, first lung function measurement and their common causes, including baseline and time-varying confounders of the mediator-outcome relationship that are also affected by exposure, and latent variables, which are assumed present but not observed.

Deprivation at birth as measured by IMD is multi-factorial and determined by a number of historical factors, and it is difficult to conceive of a single intervention (specifically one that would satisfy the consistency condition^23–25^) that could plausibly be implemented for subsets of the CF population and could shift this exposure. However, one can conceive of interventions that could be implemented to influence weight gain for children with CF, which is down-stream of the effect of deprivation at birth and also related to lung function at around age six. This motivates defining our effects of interest in terms of interventional disparity effects which are a variant of interventional effects^26,27^. They are based on the ideas of counterfactual disparity measures by VanderWeele & Robinson^28^ and Naimi et al^29^ and were formally defined in the context of multiple mediators by Micali et al^14^. Interventional disparity effect measures define the indirect effects in terms of the effect on the expected outcome in the exposed group if we could intervene on a mediator so as to shift the mediator distribution in the exposed to that in the unexposed; direct effects are defined in terms of the association that would remain between exposure and expected outcome if we could intervene on a mediator so as to shift the mediator distribution in the exposed to that in the unexposed. In our context, this enables us to estimate how the association between deprivation at birth and lung function at around age six would change if we could intervene on weight trajectory up to age six. In contrast to interventional effects, interventional disparity measures do not give a causal interpretation to the effect of the exposure on the outcome.

We further extend this approach by considering a hypothetical intervention on a longitudinal mediator trajectory. This is similar to the extension of interventional effects to incorporate time-varying exposures and mediators^30,31^.

Let *X* be the exposure of interest, here SECs at birth. We are interested in the contrast between those born in the least and most deprived quintiles of the population; let *X = 0* indicate that children were born in the least deprived quintile and *X = 1* indicate that children were born in the most deprived quintile. Let *Y* be the outcome (first lung function measurement at age [6-9)) and ***C*** the vector of baseline covariates (sex, year of birth, genotype). Let *M*_*t*_ be a time-varying mediator (weight) and *L*_*t*_ a time-varying confounder of the mediator-outcome relationship affected by the exposure (infection), where t denotes age. The complete set of mediator measurements up to age T, when the outcome is measured, is denoted 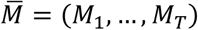 and similarly 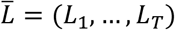. Let 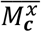 denote a random draw from the distribution of mediator trajectories in those where *X=x* and ***C****=****c***. For justification of conditioning the draws on ***C***, see Supplementary Material. The *interventional disparity measure direct effect* (IDM-DE) is the difference in the expected lung function between children from the least and most deprived quintiles, if the weight trajectories of all children were randomly drawn from the distribution of those observed in the least deprived children within strata of baseline confounders, standardized to the distribution of baseline confounders across both groups of children. Formally:

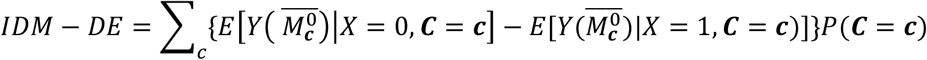

The *interventional disparity measure indirect effect (IDM-IE)* is the difference in expected lung function amongst the most deprived children if we were to randomly draw their weight trajectories from the distribution of these in least deprived children versus if we were to randomly draw their weight trajectories from the distribution of these in the most deprived children, again standardized to the distribution of baseline confounders across both groups of children. Formally:

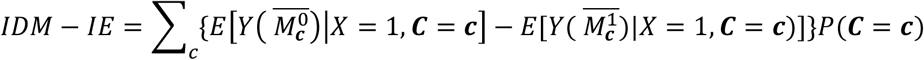

Finally, the *total adjusted association (Adj-TA)* is the difference in expected lung function between the least and most deprived children standardized to the distribution of baseline confounders across the two groups of children. Formally:

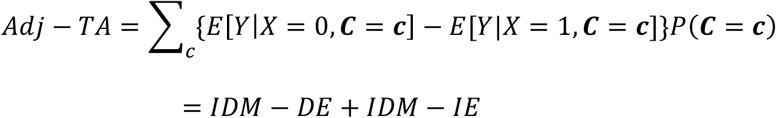

In all the estimands corresponding integrals and densities are used for continuous **C**.

The IDM-IE captures the entire effect of deprivation on lung function via weight including any effect from deprivation to infection to weight to lung function. However, it does not include any effect of deprivation on lung function via infection if this is not also mediated by weight. For justification of this see Supplementary Material.

#### Assumptions required for estimation using registry data

We estimate the estimands specified above using UK CF Registry data. This requires several assumptions: 1. no interference 2. consistency and 3. no unmeasured confounding of the mediator-outcome relationship. For explanations of assumptions 1. and 2. in this context, see Supplementary Material. The third assumption of ‘no unmeasured confounding of the mediator-outcome relationship’ includes both baseline and time-dependent confounders. We include sex, genotype and year of birth as baseline mediator-outcome confounders, and *P. aeruginosa* infection as a time-dependent confounder. However, we are not able to control for development in utero, which we believe could confound the relationship between weight and lung function (Figure 1) but which we do not have proxy measures of in the registry data. We will assess the implications of this in the discussion.

As we do not aim to infer a causal effect of the exposure on the outcome, the assumptions required for identifiability of these effects are weaker than for other causal mediation estimands. In particular, we do not have to assume no unmeasured confounding of the exposure-outcome relationship.

Assuming all 3 conditions are met, we can identify the effects of interest from the observed data (Supplementary Material).

#### Estimation

Full details of the estimation procedure are given in the Supplementary Material. We provide a brief overview here. We discretized time and assumed that one weight and infection measure per year were recorded as is the guideline in the UK. Our discrete time intervals ran from one birthday up to but not including the next birthday. For example, measurements recorded after a child’s sixth birthday but before they turned seven, were labelled as measurements at age 6. We used Monte Carlo simulation to estimate the effects of interest^14,27^. This required a number of parametric models to be specified: (1) for the expected outcome *Y* conditional on exposure, mediator at ages 0-6, baseline confounders and time-varying intermediate confounders at ages 0-6; (2) for the distribution of the intermediate confounder *L*_*t*_ at each age 0-6 conditional on exposure, baseline confounders, mediator value at the previous age and previous intermediate confounder value; and (3) for the distribution of the mediator *M*_*t*_ at ages 0-6 conditional on exposure, previous mediator value, baseline confounders and intermediate confounder value at the same age. These models were fitted to data from all children in the study. We used non-parametric bootstrap (1,000 bootstrap samples) for variance estimation. To reduce Monte-Carlo error, all the simulations were performed on a 1,200-time expanded dataset, where the 1,200 was chosen to render negligible the Monte-Carlo error in the results at the chosen number of decimal places.

#### Duplicate entries and missing data

In some cases, children had multiple measurements in a year, for example if they moved between clinics. In these cases, we selected the data from the visit closest to the child’s next birthday that had complete data on weight and infection status.

We used multiple multivariate imputation by chained equations^32^ to impute data if weight and infection status were missing for any years (10 imputed datasets). We applied the estimation algorithm to each imputed dataset and pooled point estimates across all 10 datasets. To estimate the 95% confidence intervals, we applied standard multiple imputation procedures^33^ using the variance estimated across bootstrap samples as approximation of the within imputation variance^34^. See supplementary materials for full details.

### Sensitivity Analyses

Lung function is measured with considerable error, due to natural day-to-day variation and measurement error. In a sensitivity analysis we used all available lung function data up to age 12 for individuals in the study population and fitted a linear mixed effects model with random intercept and random slope adjusted for sex, genotype, year of birth and deprivation quintile. Based on this model we predicted lung function at age six for all individuals which we used as an alternative outcome.

Due to the relatively small sample size, the parametric models specified for the estimation procedure did not include interaction terms or higher order terms but we conducted a sensitivity analysis, where we included interaction terms between deprivation and all covariates in the parametric models (1)-(3) described above.

## Results

### Study population

853 children met the eligibility criteria (Figure 2) including 165 and 172 children in the least and most deprived quintiles respectively (Table 1).

**Table 1:** Demographics and clinical characteristics of the study population stratified by deprivation quintile.

|  | 1=least deprived | 2 | 3 | 4 | 5=most deprived | Overall |
| --- | --- | --- | --- | --- | --- | --- |
| n | 165 | 164 | 180 | 172 | 172 | 853 |
| Male (%) | 95 (57.6) | 76 (46.3) | 89 (49.4) | 79 (45.9) | 94 (54.7) | 433 (50.8) |
| F508 class (%) |  |  |  |  |  |  |
| Heterozygous | 72 (43.6) | 74 (45.1) | 72 (40.0) | 67 (39.0) | 67 (39.0) | 352 (41.3) |
| Homozygous | 77 (46.7) | 74 (45.1) | 91 (50.6) | 90 (52.3) | 86 (50.0) | 418 (49.0) |
| other | 16 ( 9.7) | 16 ( 9.8) | 17 ( 9.4) | 15 ( 8.7) | 19 (11.0) | 83 ( 9.7) |
| Cohort 2007-2010 (%) | 107 (64.8) | 110 (67.1) | 120 (66.7) | 115 (66.9) | 115 (66.9) | 567 (66.5) |
| First measured lung function mean (SD*) | 93.11 (13.87) | 91.39 (14.01) | 90.91 (14.97) | 89.04 (16.68) | 88.56 (17.57) | 90.58 (15.56) |
| Age at first lung function measure (%) |  |  |  |  |  |  |
| 6 | 138 (83.6) | 142 (86.6) | 152 (84.4) | 136 (79.1) | 143 (83.1) | 711 (83.4) |
| 7 | 24 (14.5) | 18 (11.0) | 23 (12.8) | 32 (18.6) | 25 (14.5) | 122 (14.3) |
| 8 | 3 ( 1.8) | 4 ( 2.4) | 5 ( 2.8) | 4 ( 2.3) | 4 ( 2.3) | 20 ( 2.3) |
| Ever diagnosed with pseudomonas (%) | 98 (59.4) | 105 (64.0) | 108 (60.0) | 101 (58.7) | 95 (55.2) | 507 (59.4) |
| Age at first pseudomonas diagnosis mean (SD) | 2.77 (1.57) | 3.23 (1.72) | 3.00 (1.80) | 2.97 (1.67) | 2.94 (1.66) | 2.99 (1.69) |
| Number of years with missing weight data (%) |  |  |  |  |  |  |
| 0 | 25 (15.2) | 25 (15.2) | 30 (16.7) | 31 (18.0) | 28 (16.3) | 139 (16.3) |
| 1 | 65 (39.4) | 62 (37.8) | 60 (33.3) | 56 (32.6) | 61 (35.5) | 304 (35.6) |
| 2 | 44 (26.7) | 51 (31.1) | 38 (21.1) | 41 (23.8) | 43 (25.0) | 217 (25.4) |
| 3 | 20 (12.1) | 12 ( 7.3) | 27 (15.0) | 24 (14.0) | 23 (13.4) | 106 (12.4) |
| 4-6 | 11 ( 6.7) | 14 ( 8.5) | 25 (13.9) | 20 (11.6) | 17 ( 9.9) | 87 (10.2) |
| Weight z-scores mean (SD) |  |  |  |  |  |  |
| at age [0 - 1) | -0.35 (1.17) | -0.54 (1.52) | -0.50 (1.47) | -0.45 (1.39) | -0.66 (1.37) | -0.49 (1.38) |
| at age [1 - 2) | 0.32 (0.88) | 0.40 (1.05) | 0.40 (1.01) | 0.27 (0.89) | 0.08 (1.14) | 0.30 (1.01) |
| at age [2 - 3) | 0.10 (0.92) | 0.26 (0.95) | 0.37 (0.92) | 0.17 (0.88) | 0.14 (1.11) | 0.21 (0.96) |
| at age [3 - 4) | 0.11 (0.92) | 0.11 (0.97) | 0.18 (0.97) | 0.09 (0.94) | -0.04 (1.07) | 0.09 (0.97) |
| at age [4 - 5) | -0.17 (0.97) | -0.00 (1.08) | 0.03 (1.02) | -0.07 (0.93) | -0.17 (1.12) | -0.08 (1.03) |
| at age [5 - 6) | -0.07 (0.95) | -0.08 (1.04) | -0.03 (0.98) | -0.15 (1.03) | -0.14 (1.11) | -0.09 (1.02) |
| at age [6 - 7) | -0.08 (0.96) | 0.01 (1.12) | -0.00 (0.98) | -0.03 (1.02) | -0.18 (1.18) | -0.06 (1.05) |

\* Standard deviation.

**Figure 2:**
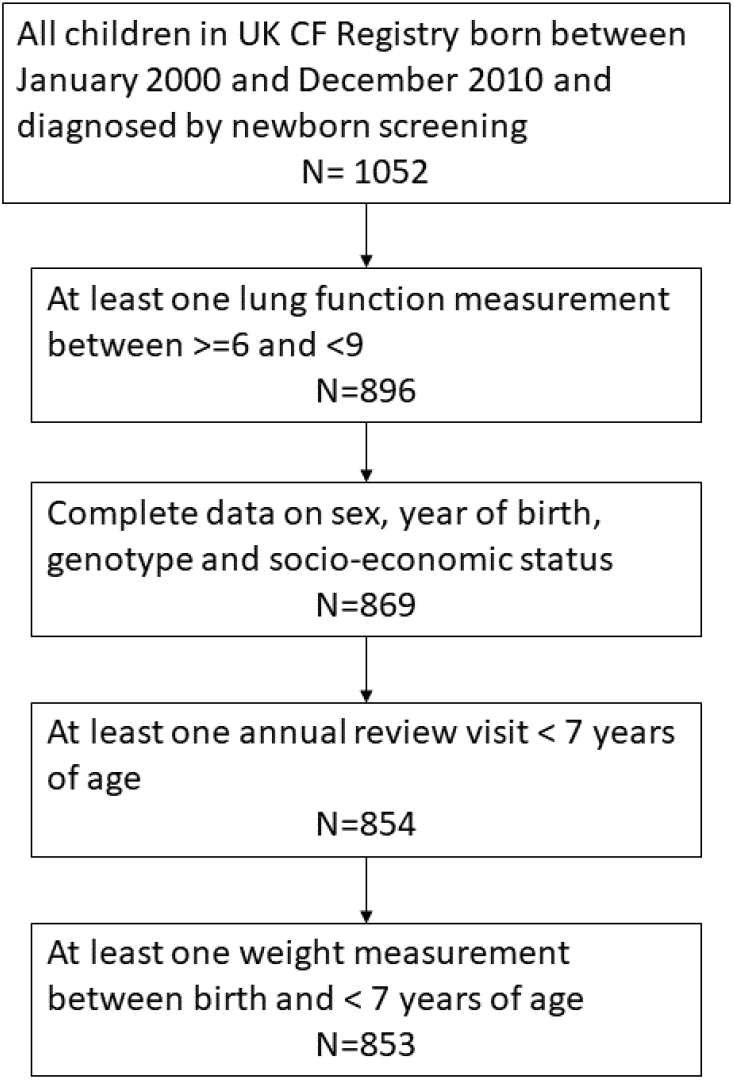
Derivation of the study population

695 individuals had at least one year from age 0-6 without a review visit, including 135 individuals in the least, and 139 individuals in the most deprived quintile (Table 1). An additional 87 annual review visits did not record weight, including 18 review visits of children in the least deprived quintile and 17 review visits of children in the most deprived quintile. Amongst the observed data, children in the most deprived quintile were on average slightly lighter than those in the least deprived quintile across almost all ages 0-6 (Table 1, Figure 3). The proportion of children ever diagnosed with *Pseudomonas* was similar across deprivation quintiles; the same was the case for mean age at first infection.

**Figure 3:**
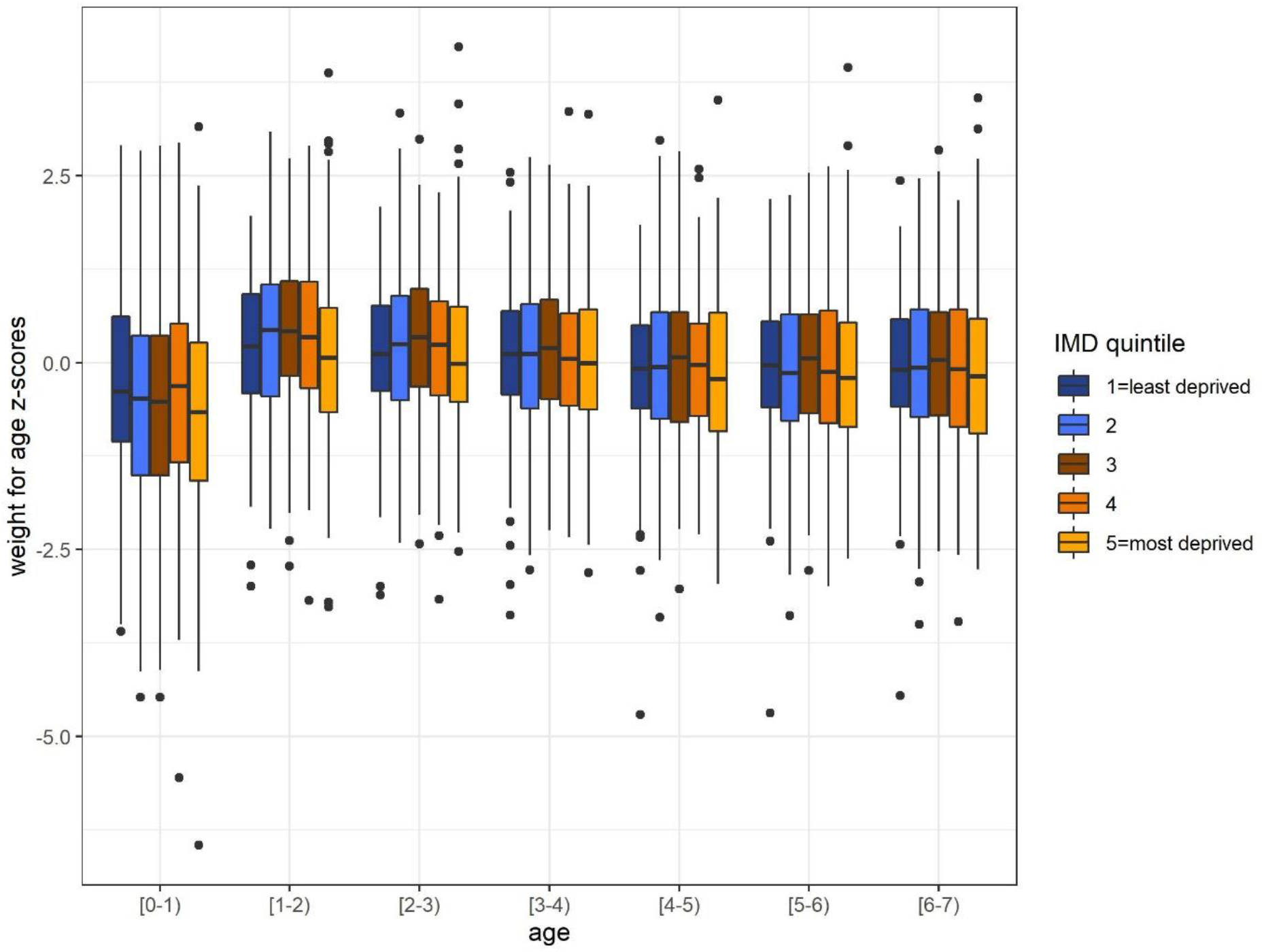
Weight for age z-scores by age as given in Table 1.

There was a social gradient in lung function at first measurement; mean lung function was 93.11 %FEV1 (SD: 13.87) and 88.56 %FEV1 (SD: 17.57) in the least and most deprived quintiles, respectively (mean difference 4.55). A Welch two sample t-test showed that this is a statistically significant difference (95% confidence interval for difference: 1.16–7.9, p-value<0.01).

### Estimated total adjusted association and interventional disparity effects

The estimated total adjusted association (Adj-TA), given as the average difference in lung function between the least and most deprived quintile of children, was 4.51 percentage points (95% CI: 1.08-7.93) (Table 2). The estimated interventional disparity measure indirect effect (IDM-IE) via weight trajectories at ages 0-6 was 0.53 percentage points (95% CI: -0.64-1.71). This is the estimated amount by which lung function would increase on average in the most deprived children if their early weight could be improved to have the same distribution as that of the least deprived children. The estimated interventional disparity measure direct effect (IDM-DE) was 3.97 percentage points (95% CI: 0.57–7.38). The difference in lung function at first measurement between the least and the most deprived children would therefore be on average 3.97 percentage points (95% CI: 0.57–7.38) even if the weight at ages 0-6 was comparable between the least and the most deprived quintiles of children.

**Table 2:** Estimated total adjusted association and interventional disparity measure direct and indirect effects. In sensitivity analysis 1 we used predicted lung function at age 6 rather than observed lung function as outcome; in sensitivity analysis 2 we included interaction terms between deprivation and all covariates in the models for weight, infection and lung function.

|  | Main analysis | Sensitivity analysis 1 | Sensitivity analysis 2 |
| --- | --- | --- | --- |
|  | Point estimate (95% confidence interval) |  |  |
| Total adjusted association | 4.51 (1.08 – 7.93) | 4.32 (1.67 – 6.98) | 4.40 (0.95 – 7.85) |
| Interventional disparity measure indirect effect (IDM-IE) | 0.53 (-0.64 – 1.71) | 0.47 (-0.46 – 1.41) | 0.23 (-1.60 – 2.05) |
| Interventional disparity measure direct effect (IDM-DE) | 3.97 (0.57 – 7.38) | 3.85 (1.24 – 6.45) | 4.17 (0.43 – 7.92) |

### Sensitivity analyses

The Adj-TA, IDM-DE and IDM-IE from the sensitivity analyses are given in Table 2. The results are comparable to the main results.

## Discussion

We assessed the relationship between socio-economic circumstances at birth and lung function at first measurement in children with CF in the UK, and the extent to which early inequalities in lung function can be explained by differences in weight trajectories from birth. Using modern methods for causal inference in longitudinal observational data, we estimated that inequalities in early lung function would only be marginally reduced (3.97 %FEV1 (95% CI: 0.57–7.38) versus 4.51 %FEV1 (95% CI: 1.08-7.93)) if early weight trajectories could be improved for the most disadvantaged children.

To address this question, we extended the framework of interventional disparity effects to the setting of a longitudinal mediator and time-varying intermediate confounder. This allowed us to separate the association between deprivation and lung function into the effects via distinct, longitudinally measured pathways, but without giving a causal interpretation to the effect of the exposure. It therefore requires weaker assumptions than more commonly used mediation methods, and may help gain insights in other areas of health inequalities research and social epidemiology where the consistency and no-unmeasured confounding (of the exposure and mediator and or/outcome) assumptions are often not satisfied^29,35^.

### Strength and Limitations

A strength of this study is that it uses the UK CF Registry data which collects data on 99% of the current CF population. We excluded just under 200 children because of missing data; the majority (156) had no lung function measurement primarily because their first lung function measurement had not yet been taken at the end of our follow-up in 2016. Other reasons for missing data were incomplete recordings of postcode which meant that deprivation data could not be linked, and no annual review data before age 7. As the main source of missingness was missing of the outcome by 2016, we are confident that the exclusion of individuals with missing data did not lead to bias and that our study population is representative of the UK CF population of children born 2000-2010.

The data used in this study are recorded systematically at clinical review visits for the purposes of reporting and research and are therefore more reliable than many observational datasets. Measurement error in lung function is, however, nevertheless likely to be non-negligible. This error is non-differential and would therefore not be expected to lead to biased estimates^36^. We also conducted a sensitivity analysis where we used longitudinally collected lung function data to get an estimate of the error-free measure at age six. As expected, the results agreed with the results from our main analysis.

The major limitation of this study is that our results rely on untestable assumptions, namely, “no interference”, “consistency” and “no unmeasured confounding of the mediator-outcome relationship”. In our setting the “no interference” assumption states that the lung function of one child is not affected by the weight trajectory of another. This is plausible especially as children with CF are encouraged to not meet in person to prevent cross-infections. The second assumption, consistency, makes assumptions about the hypothetical intervention that would shift the weight distribution in the most deprived children. We can imagine here a potentially complex intervention that could combine financial support with aspects of higher levels of training, engagement and support from dieticians for children growing up in disadvantaged circumstances and their families. The consistency assumption states that this intervention would not change the outcome for those children whose weight trajectory was set to the trajectory that was actually observed. However, it is conceivable that stronger engagement and more support for more deprived families would improve several other factors which may lead to improved lung function, such as clinic attendance and self-confidence in other aspects of CF care as well as reduced parental stress and improved mental health. Therefore, although we did not find any evidence that a hypothetical intervention on weight would reduce inequalities in lung function significantly, it may have positive effects beyond any direct effects on weight. Finally, we had to assume no unmeasured confounding of the mediator-outcome relationship. According to our assumed causal relationships (Figure 1), it is plausible that development in-utero impacts early weight and lung development and there is some evidence that supports that birthweight (but not gestational age) is associated with lung function at age six^37,38^. Unfortunately, the CF registry has only recently started collecting data on birthweight and there was no proxy in the dataset that could be used to assess the impact of ignoring development in-utero in our analysis. However, as the effect of growth in-utero would have positive effects on early growth and lung function, then any hypothetical intervention on weight after birth would likely have even less of an impact than our estimated effect.

Our analysis further relies on the correct specification of the regression models used in the estimation procedure. Due to the small sample size, we did not include interactions and higher order terms in our models for the main analysis. However, we did conduct a sensitivity analysis in which we included interactions between deprivation and all covariates in the models. This did not substantially change our results although the uncertainty around estimates was increased.

### Comparison with other studies and implications for further research and clinical practice

Weight in the first years of life has been shown to be strongly associated with later lung function in children with CF^1,2,39–42^ which has led to the development of CF clinical practice guidelines for nutrition management^43^. However, this is the first study that we are aware of that has looked at the role of early weight in the generation of socio-economic inequalities in lung function in CF. Although optimal nutrition is undoubtedly important, we found that reducing inequalities in early weight would likely only have a marginal effect on inequalities in later lung function.

Other factors that may lie on the pathway from SECs to lung function have previously been studied, including the prescription of antibiotics^3^, exposure to environmental tobacco smoke^44,45^, and age at diagnosis^5^. However, it remains unclear what drives inequalities in early lung function and further research is needed. There is anecdotal evidence that the regularity of clinic attendance may display a social pattern due to barriers that more deprived families face regarding transportation and the ability to take time off work. Also, engagement with the clinical care team and health literacy of parents from more disadvantaged backgrounds may be lower than in more advantaged families and may have a role to play^46,47^. In addition, environmental factors and living conditions need further consideration.

## Conclusion

There is a strong association between socio-economic conditions at birth and early lung function for children with CF which we estimated would only be marginally reduced if early weight trajectories could be improved for the most disadvantaged children. Further research is needed to identify modifiable pathways to inequalities for children with CF such that the underlying factors can be addressed. In the meantime, action to address poverty and improve social conditions for disadvantaged children growing up with CF is required to address health inequalities.

The methods developed and applied here may also help to give insights into the generation of inequalities in other conditions.

## Data Availability

The data used in this study are from the UK CF Registry and are available from the UK CF Registry on reasonable request.

## Acknowledgements

We thank the UK CF Registry team, and the UK CF centres and clinics for submitting data to the Registry. Special thanks to the people with cystic fibrosis and their families who have agreed for their UK CF Registry data to be used for research.

