## Supplementary Material for "How do growth and nutrition explain social inequalities in lung function in children with cystic fibrosis? A longitudinal mediation analysis using interventional disparity effects with time-varying mediators and intermediate confounders"

Daniela K. Schlueter, Ruth H. Keogh, Rhian Daniel, Shadrac Agbla, David Taylor-Robinson

### S1. Additional information on the rationale of the effects of interest

Genotype, sex and year of birth influence weight in children with CF. In practice a hypothetical intervention to improve weight would be applied in the same way to all children from socio-economically disadvantaged backgrounds. Therefore it may be impossible to expect that, for example, the distribution of weight in children who are homozygous F508del could be the same as that of children who are heterozygous F508del. To reflect this in our analysis, we restrict the draws from the distribution of the mediator to be within strata of these confounders.

The relationship between weight and lung function is confounded by infection status which itself is affected by deprivation and weight (Figure 1 in the main text). Similarly as we argue above, a dependence between weight and infection trajectories would likely remain following a hypothetical invervention that is applied to all children from disadvantaged backgrounds in the same way across ages. However, we cannot estimate the effect of this kind of intervention using the methods applied here. Instead, our hypothetical intervention removes the dependence between weight and infection. Practically, we can think about this as being a responsive intervention - if a child has had an infection the intervention applied to them is 'intensified' in some way to break the dependence of weight on infection. This is not an unreasonable scenario as children receiving the intervention would still be monitored and while clinicians know the difference between the expected ditributions of weight depending on sex, genotype, year of birth and SES and may therefore not aim to break the dependence between weight and the baseline confounders, it would not be possible for a clinician to disentangle any effects of deprivation on weight directly from any effects on weight via earlier infection. Therefore, a realistic intervention could aim for similar weight distributions between least and most deprived groups stratified by sex, year of birth and genotype but not take into account infection. Our estimands therefore capture the entire effect of deprivation on lung function via weight including any effect from deprivation to infection to weight to lung function. However, it does not include any effect of deprivation on lung function via infection if this is not mediated by weight.

It has been pointed out that this approach may not be suitable to give insights into the mechanisms by which SES affects lung function [1]. However, as outlined above, we believe that our estimands address a realistic practical question of the pulmonary effect of a hypothetical intervention on weight which was the focus of this study.

### S2. Assumptions required for estimation using registry data

We estimate the estimands specified in the main text using UK CF Registry data. This requires several assumptions: 1. no interference 2. consistency and 3. no unmeasured confounding of the mediator-outcome relationship. No interference in this context means that the lung function of one child is not affected by the weight trajectory of another. Consider a group of children with the same exposure, same baseline confounders, same infection status trajectory and same weight trajectory, say $\bar{m}$. The consistency assumption in our application states that if we were to set the weight trajectory in this group to $\bar{m}$, the mean lung function (at age six) would be the same as the observed lung function in that group. This assumption therefore states that the hypothetical intervention does not have any effect on lung function via any pathways other than weight and that the effect of setting the weight trajectory to a particular set of values via our hypothesized intervention is the same as if the same weight trajectory were obtained naturally.

The third assumption of 'no unmeasured confounding of the mediator-outcome relationship' includes both baseline and time-dependent confounders. We include sex, genotype and year of birth as baseline mediator-outcome confounders, and P. aeruginosa infection as a time-dependent confounder. However, we are not able to control for development in utero, which we believe could confound the relationship between weight and lung function (see Figure 1 in the main text) but which we do not have proxy measures of in the registry data. We will assess the implications of this in the discussion in the main text. As we do not aim to infer a causal effect of the exposure on the outcome, the assumptions required for identifiability of these effects are weaker than for other causal mediation estimands. In particular we do not have to assume no unmeasured confounders of the exposure- outcome relationship. Assuming all 3 conditions are met, we can identify the effects of interest from the observed data. Details of how identification is achieved are outlined below.

### S3. Identifiability

Let $X$ be the exposure of interest, here socio-economic conditions at birth. We are interested in the contrast between those born in the least and most deprived quintiles of the population. Therefore, let $X = 0$ indicate that children were born in the least deprived quintile and $X = 1$ indicate that children were born in the most deprived quintile. Let $Y$ be the outcome (first lung function measurement at age [6-9)), $\mathbf{C}$ a vector of possible baseline confounders (sex, year of birth, genotype). Let $M_t$ be a time-varying mediator (weight) and $L_t$ a time-varying confounder of the mediator-outcome relationship affected by the exposure (infection). For ease of presentation we initially consider the mediator and time-varying confounder at 2 time points only. Let $Y(m_1, m_2)$ denote the potential value that Y would take if $M_1$ and $M_2$ were intervened upon and set to $m_1$ and $m_2$. Finally let $\bar{M}_c^x$ denote a random draw from the distribution of mediator trajectories (over the two time points) in those where $X = x$ and $\mathbf{C} = \mathbf{c}$.

The assumed causal relationship between the variables is given in Supplementary Figure 1.

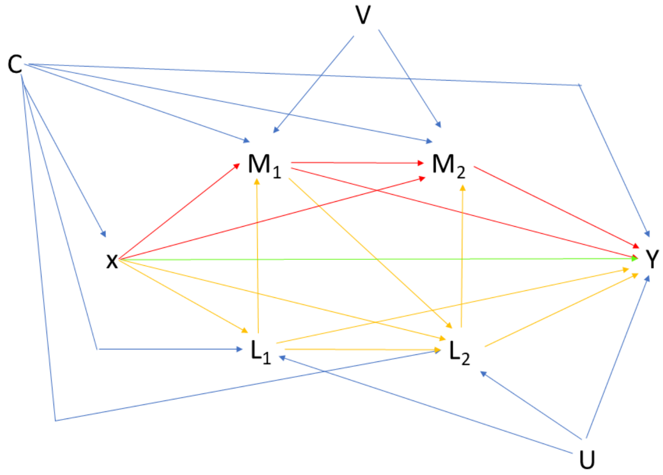


Supplementary Figure 1: Assumed causal relationship between variables where X is the exposure (socio-economic status), $M_1$ and $M_2$ the mediator (weight) at time points 1 and 2, $L_1$ and $L_2$ the time varying confounder of the mediator outcome relationship affected by exposure (infection), Y the outcome (lung function), C the measured baseline confounders (sex, year of birth, genotype) and U and V unmeasured confounders. The colours of the arrows are for readability only.

The interventional disparity measure direct and indirect effects (defined here for discrete C but adaptable to continuous covariates by replacement of sums by integrals and probabilities by densities), are given by

$$\text{IDM-DE} = \sum_c \{E[Y(\bar{M}_{\mathbf{c}}^0)|X = 0, \mathbf{C} = \mathbf{c})] - E[Y(\bar{M}_{\mathbf{c}}^0)|X = 1, \mathbf{C} = \mathbf{c})]\} \Pr(\mathbf{C} = \mathbf{c}) \quad (1)$$

and

$$\text{IDM-IE} = \sum_c \{E[Y(\bar{M}_{\mathbf{c}}^0)|X = 1, \mathbf{C} = \mathbf{c})] - E[Y(\bar{M}_{\mathbf{c}}^1)|X = 1, \mathbf{C} = \mathbf{c})]\} \Pr(\mathbf{C} = \mathbf{c}) \quad (2)$$

respectively.

Identifiability of these effects therefore requires the identifiability of $E[Y(\bar{M}_{\mathbf{c}}^{x_1})|X = x_2, \mathbf{C} = \mathbf{c})]$ for $x_1 = 0, 1$ and $x_2 = 0, 1$. We can write this probability as follows, where changes made in each line are shown in red:

$E[Y(\bar{M}_{\mathbf{c}}^{x_1})|X = x_2, \mathbf{C} = \mathbf{c})] =$

by definition of interventional effect

$$= \sum_{m_1,m_2} \{E[Y(m_1, m_2)|X = x_2, \mathbf{C} = \mathbf{c}] \times \Pr(M_1 = m_1, M_2 = m_2|X = x_1, \mathbf{C} = \mathbf{c})\}$$

by law of total expectation

$$= \sum_{m_1,m_2} \sum_{l_1} \{E[Y(m_1, m_2)|X = x_2, \mathbf{C} = \mathbf{c}, L_1 = l_1] \times \Pr(L_1 = l_1|X = x_2, \mathbf{C} = \mathbf{c}) \times \Pr(M_1 = m_1, M_2 = m_2|X = x_1, \mathbf{C} = \mathbf{c})\}$$

by conditional exchangability assumption

$$= \sum_{m_1,m_2} \sum_{l_1} \{E[Y(m_1, m_2)|X = x_2, \mathbf{C} = \mathbf{c}, L_1 = l_1, M_1 = m_1] \times \Pr(L_1 = l_1|X = x_2, \mathbf{C} = \mathbf{c}) \times \Pr(M_1 = m_1, M_2 = m_2|X = x_1, \mathbf{C} = \mathbf{c})\}$$

by law of total expectation

$$= \sum_{m_1,m_2} \sum_{l_1,l_2} \{E[Y(m_1, m_2)|X = x_2, \mathbf{C} = \mathbf{c}, L_1 = l_1, L_2 = l_2, M_1 = m_1] \times \Pr(L_2 = l_2|L_1 = l_1, M_1 = m_1, X = x_2, \mathbf{C} = \mathbf{c}) \times \Pr(L_1 = l_1|X = x_2, \mathbf{C} = \mathbf{c}) \times \Pr(M_1 = m_1, M_2 = m_2|X = x_1, \mathbf{C} = \mathbf{c})\}$$

by conditional exchangability assumption

$$= \sum_{m_1,m_2} \sum_{l_1,l_2} \{E[Y(m_1, m_2)|X = x_2, \mathbf{C} = \mathbf{c}, L_1 = l_1, L_2 = l_2, M_1 = m_1, M_2 = m_2] \times \Pr(L_2 = l_2|L_1 = l_1, M_1 = m_1, X = x_2, \mathbf{C} = \mathbf{c}) \times \Pr(L_1 = l_1|X = x_2, \mathbf{C} = \mathbf{c}) \times \Pr(M_1 = m_1, M_2 = m_2|X = x_1, \mathbf{C} = \mathbf{c})\}$$

by consistency assumption

$$= \sum_{m_1,m_2} \sum_{l_1,l_2} \{E[Y|X = x_2, \mathbf{C} = \mathbf{c}, L_1 = l_1, L_2 = l_2, M_1 = m_1, M_2 = m_2] \times \Pr(L_2 = l_2|L_1 = l_1, M_1 = m_1, X = x_2, \mathbf{C} = \mathbf{c}) \times \Pr(L_1 = l_1|X = x_2, \mathbf{C} = \mathbf{c}) \times \Pr(M_1 = m_1, M_2 = m_2|X = x_1, \mathbf{C} = \mathbf{c})\}$$

Similarly, it can be shown that for any number of time points $T$

$$
\begin{aligned}
E[Y(\bar{M}_c^{x_1})|X=x_2,\mathbf{C}=\mathbf{c})] = &\sum_{\bar{m}}\sum_{\bar{l}}\{E[Y|X=x_2,\mathbf{C}=\mathbf{c},\bar{L}=\bar{l},\bar{M}=\bar{m}] \\
&\times\prod_{t=1}^{T}\Pr(L_t=l_t|X=x_2,\mathbf{C}=\mathbf{c},L_{t-1}=l_{t-1},M_{t-1}=m_{t-1}) \\
&\times\prod_{t=1}^{T}\Pr(M_t=m_t|X=x_1,\mathbf{C}=\mathbf{c},M_{t-1}=m_{t-1})
\end{aligned}
$$

In this expression we can also specify the distribution of $M_1$,$M_2$,..,$M_t$ conditional on L and then marginalised out $L$:

$$
\begin{aligned}
E[Y(\bar{M}_c^{x_1})|X=x_2,\mathbf{C}=\mathbf{c})] = &\sum_{\bar{m}}\sum_{\bar{l}}\{E[Y|X=x_2,\mathbf{C}=\mathbf{c},\bar{L}=\bar{l},\bar{M}=\bar{m}] \\
&\times\prod_{t=1}^{T}\Pr(L_t=l_t|X=x_2,\mathbf{C}=\mathbf{c},L_{t-1}=l_{t-1},M_{t-1}=m_{t-1}) \\
&\times\sum_{\bar{l}'}\prod_{t=1}^{T}\{\Pr(M_t=m_t|X=x_1,\mathbf{C}=\mathbf{c},M_{t-1}=m_{t-1},L_t=l'_t) \\
&\times\Pr(L_t=l'_t|X=x_1,\mathbf{C}=\mathbf{c},M_{t-1}=m_{t-1},L_{t-1}=l'_{t-1})\}
\end{aligned}
$$

This is the approach we have taken in our analysis as specified below.

Therefore identifiability is given for interventional disparity direct and indirect effects which consider $T$ measurements of mediator and intermedate confounders.

### S4. Estimation by Monte Carlo simulation using complete data

*Using complete data on the whole population*:

1. Fit linear models to observed data for weight at each annual review visit $t$ ($M_t$) condtional on exposure X, baseline confounders $\mathbf{C}$, infection during the preceding year which is assessed at the current visit ($L_t$), and weight at the previous visit ($M_{t-1}$) as outlined in the DAG. $M_0$ (weight up to the first birthday) is regressed on exposure X, baseline confounders $\mathbf{C}$ and infection assessed at the current visit visit ($L_0$) only.

$$M_t = \alpha_0 + \alpha_1 X + \boldsymbol{\alpha_2 C} + \alpha_3 M_{t-1} + \alpha_4 L_t + \epsilon_M \tag{3}$$

and

$$\epsilon_M \sim N(0, \sigma_M^2) \tag{4}$$

2. Fit logistic regression models to observed data for infection recorded at each annual review visit $t$ conditional on exposure X, baseline confounders $\mathbf{C}$, infection status at the previous visit ($L_{t-1}$), and weight at the previous visit ($M_{t-1}$) as outlined in the DAG. $L_0$ (infection up to the first birthday) is regressed on exposure X, and baseline confounders $\mathbf{C}$ only.

$$\log\left(\frac{p_{l_t}|X,\mathbf{C},M_{t-1},L_{t-1}}{(1-p_{l_t}|X,\mathbf{C},M_{t-1},L_{t-1})}\right) = \beta_0 + \beta_1 X + \boldsymbol{\beta_2 C} + \beta_3 M_{t-1} + \beta_4 L_{t-1} \tag{5}$$

and

$$L_t \sim Binom(p_{l_t}|X, \mathbf{C}, M_{t-1}, L_{t-1}) \tag{6}$$

3. Fit linear model for the expected outcome Y (first lung function measurement) conditional on exposure X, baseline confounders **C**, weight at all ages 0-6 ($\bar{M}$) and infection status at all ages 0-6 ($\bar{L}$) as outlined in the DAG

$$E[Y|X, \boldsymbol{C}, \bar{\boldsymbol{M}}, \bar{\boldsymbol{L}}] = \gamma_0 + \gamma_1 X + \boldsymbol{\gamma_2 C} + \boldsymbol{\gamma_3 \bar{M}} + \boldsymbol{\gamma_4 \bar{L}} \tag{7}$$

*Using data from the individuals in the least and most deprived groups (unexposed and exposed) only*:

4. Expand the data set N times; to reduce Monte-Carlo error, all the simulations were performed on an N=1,200-time expanded dataset, where the 1,200 was chosen to render negligible the Monte-Carlo error in the results at the chosen number of decimal places.
5. For each individual $i$ in each row in the expanded dataset generate a random draw, denoted $\bar{m}^0_{ic}$, from the distribution of the mediator trajectory in the unexposed (least deprived) conditional on baseline covariates $\mathbf{C} = \mathbf{c_i}$ by
   (a) Setting exposure X to 0 (least deprived) for all individuals
   (b) For $t = 1, ..., T$ iteratively simulating weight and infection trajectories based on parameter estimates from step 1 and step 2, $\mathbf{C} = \mathbf{c_i}$, $X = 0$ and simulated previous weight and infection status. For $M_t$ this involves sampling from a normal distribution with mean and variance given by equations (3) and (4). For $L_t$ it involves sampling from a binomial distribution with probability given by equation (5).
6. For each individual $i$ in the expanded dataset generate a random draw, denoted $\bar{m}^1_{ic}$, from the distribution of the mediator trajectory in the exposed (most deprived) conditional on baseline covariates $\mathbf{C} = \mathbf{c_i}$ by
   (a) Setting exposure X to 1 (most deprived) for all individuals
   (b) For $t = 1, ..., T$ iteratively simulating weight and infection trajectories based on parameter estimates from step 1 and step 2, $\mathbf{C} = \mathbf{c_i}$, $X = 1$ and simulated previous weight and infection status.
7. Estimate standardised expected lung function among the exposed (most deprived) if their weight trajectory was randomly drawn from the distribution of the exposed (most deprived), conditional on baseline covariates $\sum_c E[Y(\bar{M}^1_c)|X = 1, \mathbf{C} = \mathbf{c}] \Pr(\mathbf{C} = \mathbf{c})$ by:
   (a) Setting exposure X to 1 (most deprived) for all individuals
   (b) Using the parameter estimates from the model for E[Y] (equation (7)), estimate the expected outcome for each individual in each row of the expanded dataset using their simulated infection trajectory $\bar{l}_i$ from step 6, the draw from the distribution of weight trajectories $\bar{m}^1_{ic}$ from step 6, their observed baseline covariates $\mathbf{c_i}$ and exposure $X = 1$.
   (c) Calculating the mean expected outcome in the expanded dataset to get an estimate for $\sum_c E[Y(\bar{M}^1_c)|X = 1, \mathbf{C} = \mathbf{c}] \Pr(\mathbf{C} = \mathbf{c})$
8. Estimate standardised expected lung function among the unexposed (least deprived) if their weight trajectory was randomly drawn from the distribution of the unexposed (least deprived), conditional on baseline covariates $\sum_c E[Y(\bar{M}^0_c)|X = 0, \mathbf{C} = \mathbf{c}] \Pr(\mathbf{C} = \mathbf{c})$ by:
   (a) Setting exposure X to 0 (least deprived) for all individuals
   (b) Using the parameter estimates from the model for E[Y] (equation (7)), estimate the expected outcome for each individual in each row of the expanded dataset using their simulated infection trajectory $\bar{l}_i$ from step 5, the draw from the distribution of weight trajectories $\bar{m}^0_{ic}$ from step 5, their observed baseline covariates $\mathbf{c_i}$ and exposure $X = 0$.
   (c) Calculating the mean expected outcome in the expanded dataset to get an estimate for $\sum_c E[Y(\bar{M}^0_c)|X = 0, \mathbf{C} = \mathbf{c}] \Pr(\mathbf{C} = \mathbf{c})$

9. Estimate standardised expected lung function among the exposed (most deprived) if their weight trajectory was randomly drawn from the distribution of the unexposed (least deprived), conditional on baseline covariates $\sum_c E[Y(\bar{M}_c^0)|X = 1, \mathbf{C} = \mathbf{c}] \Pr(\mathbf{C} = \mathbf{c})$ by:
   (a) Setting exposure X to 1 (most deprived) for all individuals
   (b) For $t = 1, ..., T$ iteratively simulating infection status based on parameter estimates from step 2 and using simulated previous infection status (t>1) $l_{t-1}$, draw from the distribution of the weight trajectories $\bar{m}_{ic}^0$ generated in step 5, observed baseline covariates $\mathbf{C} = \mathbf{c_i}$ and exposure X=1.
   (c) Using the parameter estimates for the model for $E[Y|X, \boldsymbol{C}, \bar{\boldsymbol{M}}, \bar{\boldsymbol{L}}]$ (equation (7)), estimate the expected outcome for each individual in each row of the expanded dataset using their simulated infection trajectory $\bar{l}_i$ from (b) , their draw from the distribution of weight trajectories $\bar{m}_{ic}^0$ from step 5, their observed baseline covariates $\mathbf{c_i}$ and exposure X=1.
   (d) Calculating the mean expected outcome in the expanded datset to get an estimate for $\sum_c E[Y(\bar{M}_c^0)|X = 1, \mathbf{C} = \mathbf{c}] \Pr(\mathbf{C} = \mathbf{c})$
10. Estimate the quantities of interets:
   (a) Estimate IDM-IE $= \sum_c E[Y(\bar{M}_c^0)|X = 1, \mathbf{C} = \mathbf{c}] \Pr(\mathbf{C} = \mathbf{c}) - \sum_c E[Y(\bar{M}_c^1)|X = 1, \mathbf{C} = \mathbf{c}] \Pr(\mathbf{C} = \mathbf{c})$ using the estimates from step 9 and step 7.
   (b) Estimate IDM-DE $= \sum_c E[Y(\bar{M}_c^0)|X = 0, \mathbf{C} = \mathbf{c}] \Pr(\mathbf{C} = \mathbf{c}) - \sum_c E[Y(\bar{M}_c^0)|X = 1, \mathbf{C} = \mathbf{c}] \Pr(\mathbf{C} = \mathbf{c})$ using estimates from step 7 and step 8.
   (c) Estimate the adjusted total association Adj-TA $= \sum_c E[Y(\bar{M}_c^0)|X = 0, \mathbf{C} = \mathbf{c}] \Pr(\mathbf{C} = \mathbf{c}) - \sum_c E[Y(\bar{M}_c^1)|X = 1, \mathbf{C} = \mathbf{c}] \Pr(\mathbf{C} = \mathbf{c})$ using the sum of the estimates of IDM-IE and IDM-DE.
11. To calculate the variance of the estimates, repeat step 1 to step 10 using 1000 bootstrap samples of the original data

### S5. Estimation by Monte Carlo simulation with missing data

1. Generate K imputed datasets, we choose K=10. We used multiple multivariate imputation by chained equations as implemented in the *mice* package in R. Baseline covariates, exposure, weight at all ages, infection at all ages and first lung function measurement were included as predictors in the imputation models for weight and infection status. We used a linear model for weight at all ages and logistic regression for infection status at all ages.
2. Repeat step 1 to step 11 for all K imputed datasets.
3. Calculate the pooled point estimates of the quantities of interest (IDM-IE, IDM-DE and ADJ-TA) by taking the mean of the point estimates across K imputed datasets $\hat{\theta}_{MI} = \frac{1}{K} \sum_{k=1}^{K} \hat{\theta}_k$ where $\hat{\theta}_k$ is a point estimate of the quantity of interest based on imputed dataset k.
4. To calculate the confidence intervals, let $\hat{W}$ denote within imputation variance and $\hat{V}$ the between imputation variance. Following [2], estimate the within imputation variance as the average of the variance of the bootstrap samples across the K imputed datasets $\hat{W} = \frac{1}{K} \sum_{k=1}^{K} var(\hat{\theta}_k)$. The between imputation variance is given by $\hat{V} = \frac{1}{(K-1)} \sum_{k=1}^{K} (\hat{\theta}_k - \hat{\theta}_{MI})^2$.The variance of the point estimate is then given by $var(\hat{\theta}_{MI}) = \hat{W} + \frac{(K+1)}{K} \hat{V}$. To construct confidence intervals, assume that $\frac{\hat{\theta}_{MI} - \theta}{\sqrt{var(\hat{\theta}_{MI}}}$ follows a $t_R$ distribution with $R = (K-1)(1 + \frac{(K\hat{W})}{(K+1)\hat{V}})$.

### S6. R Code

Below is the code that takes a list of 10 imputed datasets ("imputed.data") and runs through steps ref{1} to step 11 of the algorithm above.

```r
library(tidyverse)
library(formatR)

set.seed(11122020)

# for loop over the imputed datasets
for (imp in 1:10) {
    data <- imputed.data[[imp]]

    # set up data frame for saving results for current dataset
    results.boot <- as.data.frame(matrix(ncol = 6, nrow = 1001))
    colnames(results.boot) <- c("TE", "IDM_IDE", "IDM_DE", "mean.lf.md.md", "mean.lf.md.ld",
        "mean.lf.ld.ld")

    # for loop over bootstrap samples
    for (z in 1:1001) {
        if (z == 1) {
            rows = 1:nrow(data)
        } else {
            rows <- sample(nrow(data), size = nrow(data), replace = T)
        }
        i1 <- data[rows, ]
        # generate new unique ids for each row in the sample
        # ('S01CaseId_Original'' is the original person identifier which will
        # potentially occur multiple times in a bootstrap sample)
        i1 <- i1 %>%
            group_by(S01CaseId_Original) %>%
            mutate(marker = 1) %>%
            mutate(rows_n = row_number()) %>%
            ungroup() %>%
            mutate(original.id = S01CaseId_Original) %>%
            mutate(S01CaseId_Original = paste(S01CaseId_Original, ".", rows_n, sep = ""))

        # centre year of birth
        i1$yob.centered <- i1$yearofbirth - 2000

        #-----------------------------------------------------
        # Regression Models -----------------------------------
        #-----------------------------------------------------
        # outcome model (lung function) conditional on weight trajectory
        # ('weight'), infection trajectory ('pseud') and confounders
        # ('dmg_sex','F508_class','yob.centered')
        lf.model2 <- lm(lf ~ dmg_sex + F508_class + yob.centered + imd.quintile +
            weight.age0_1 + weight.age1_2 + weight.age2_3 + weight.age3_4 + weight.age4_5 +
            weight.age5_6 + weight.age6_7 + pseud.age0_1 + pseud.age1_2 + pseud.age2_3 +
            pseud.age3_4 + pseud.age4_5 + pseud.age5_6 + pseud.age6_7, data = i1)

        # models for weight at ages 0-6
        w0.model <- lm(weight.age0_1 ~ dmg_sex + F508_class + yob.centered + imd.quintile +
            pseud.age0_1, data = i1)
        w1.model <- lm(weight.age1_2 ~ dmg_sex + F508_class + yob.centered + imd.quintile +
            pseud.age1_2 + weight.age0_1, data = i1)
        w2.model <- lm(weight.age2_3 ~ dmg_sex + F508_class + yob.centered + imd.quintile +
```

```
    pseud.age2_3 + weight.age1_2, data = i1)
w3.model <- lm(weight.age3_4 ~ dmg_sex + F508_class + yob.centered + imd.quintile +
    pseud.age3_4 + weight.age2_3, data = i1)
w4.model <- lm(weight.age4_5 ~ dmg_sex + F508_class + yob.centered + imd.quintile +
    pseud.age4_5 + weight.age3_4, data = i1)
w5.model <- lm(weight.age5_6 ~ dmg_sex + F508_class + yob.centered + imd.quintile +
    pseud.age5_6 + weight.age4_5, data = i1)
w6.model <- lm(weight.age6_7 ~ dmg_sex + F508_class + yob.centered + imd.quintile +
    pseud.age6_7 + weight.age5_6, data = i1)

# models for infection status at ages 0-6
i0.model <- glm(pseud.age0_1 ~ dmg_sex + F508_class + yob.centered + imd.quintile,
    family = binomial(link = "logit"), data = i1)
i1.model <- glm(pseud.age1_2 ~ dmg_sex + F508_class + yob.centered + imd.quintile +
    weight.age0_1 + pseud.age0_1, family = binomial(link = "logit"), data = i1)
i2.model <- glm(pseud.age2_3 ~ dmg_sex + F508_class + yob.centered + imd.quintile +
    weight.age1_2 + pseud.age1_2, family = binomial(link = "logit"), data = i1)
i3.model <- glm(pseud.age3_4 ~ dmg_sex + F508_class + yob.centered + imd.quintile +
    weight.age2_3 + pseud.age2_3, family = binomial(link = "logit"), data = i1)
i4.model <- glm(pseud.age4_5 ~ dmg_sex + F508_class + yob.centered + imd.quintile +
    weight.age3_4 + pseud.age3_4, family = binomial(link = "logit"), data = i1)
i5.model <- glm(pseud.age5_6 ~ dmg_sex + F508_class + yob.centered + imd.quintile +
    weight.age4_5 + pseud.age4_5, family = binomial(link = "logit"), data = i1)
i6.model <- glm(pseud.age6_7 ~ dmg_sex + F508_class + yob.centered + imd.quintile +
    weight.age5_6 + pseud.age5_6, family = binomial(link = "logit"), data = i1)

#-----------------------------------------------------------
# Select least and most deprived groups and expand dataset--
#-----------------------------------------------------------
# select only those from the dataset that are in the most or least
# deprived quintile
i1.new <- filter(i1, imd.quintile %in% c(1, 5))
# expand dataset 1200 times to reduce monte carlo error and generate
# unique IDs for each row in the dataframe
i1.new.expanded <- i1.new %>%
    slice(rep(1:n(), each = 1200)) %>%
    group_by(S01CaseId_Original) %>%
    mutate(marker2 = 1) %>%
    mutate(rows_n2 = row_number()) %>%
    ungroup() %>%
    mutate(original.id = S01CaseId_Original) %>%
    mutate(S01CaseId_Original = paste(S01CaseId_Original, ".", rows_n2, sep = ""))

#---------------------------------------------------------------
# simulate weight and infection for exposure = most deprived ----
#---------------------------------------------------------------
# set up dataset to save results
simulated.outcomes.md <- as.data.frame(matrix(nrow = 7 * nrow(i1.new.expanded),
    ncol = 5))
colnames(simulated.outcomes.md) <- c("id", "age", "exposure", "weight", "infection")
newdata.md <- select(i1.new.expanded, S01CaseId_Original, dmg_sex, F508_class,
    yob.centered, imd.quintile)
# dataset used for simulation
```

```r
newdata.md$imd.quintile = "5"
newdata.md <- arrange(newdata.md, S01CaseId_Original)

# t=0-1
pseud0 <- rbinom(n = nrow(newdata.md), prob = predict(i0.model, newdata.md,
    type = "response"), size = 1)
simulated.outcomes.md[1:nrow(i1.new.expanded), c(1:3, 5)] <- cbind.data.frame(newdata.md$S01Cas[illegible]
    rep(0, nrow(i1.new.expanded)), rep("md", nrow(i1.new.expanded)), pseud0,
    stringsAsFactors = F)
w0 <- predict(w0.model, cbind(newdata.md, pseud.age0_1 = as.factor(pseud0))) +
    rnorm(nrow(newdata.md), mean = 0, sd = sigma(w0.model))
simulated.outcomes.md[1:nrow(i1.new.expanded), 4] <- w0

# t=1-2
pseud1 <- rbinom(n = nrow(newdata.md), prob = predict(i1.model, cbind(newdata.md,
    pseud.age0_1 = as.factor(pseud0), weight.age0_1 = w0), type = "response"),
    size = 1)
simulated.outcomes.md[(nrow(i1.new.expanded) + 1):(2 * nrow(i1.new.expanded)),
    c(1:3, 5)] <- cbind.data.frame(newdata.md$S01CaseId_Original, rep(1,
    nrow(i1.new.expanded)), rep("md", nrow(i1.new.expanded)), pseud1, stringsAsFactors = F)
w1 <- predict(w1.model, cbind(newdata.md, pseud.age1_2 = as.factor(pseud1),
    weight.age0_1 = w0)) + rnorm(nrow(newdata.md), mean = 0, sd = sigma(w1.model))
simulated.outcomes.md[(nrow(i1.new.expanded) + 1):(2 * nrow(i1.new.expanded)),
    4] <- w1

# t=2-3
pseud2 <- rbinom(n = nrow(newdata.md), prob = predict(i2.model, cbind(newdata.md,
    pseud.age1_2 = as.factor(pseud1), weight.age1_2 = w1), type = "response"),
    size = 1)
simulated.outcomes.md[(2 * nrow(i1.new.expanded) + 1):(3 * nrow(i1.new.expanded)),
    c(1:3, 5)] <- cbind.data.frame(newdata.md$S01CaseId_Original, rep(2,
    nrow(i1.new.expanded)), rep("md", nrow(i1.new.expanded)), pseud2, stringsAsFactors = F)
w2 <- predict(w2.model, cbind(newdata.md, pseud.age2_3 = as.factor(pseud2),
    weight.age1_2 = w1)) + rnorm(nrow(newdata.md), mean = 0, sd = sigma(w2.model))
simulated.outcomes.md[(2 * nrow(i1.new.expanded) + 1):(3 * nrow(i1.new.expanded)),
    4] <- w2

# t=3-4
pseud3 <- rbinom(n = nrow(newdata.md), prob = predict(i3.model, cbind(newdata.md,
    pseud.age2_3 = as.factor(pseud2), weight.age2_3 = w2), type = "response"),
    size = 1)
simulated.outcomes.md[(3 * nrow(i1.new.expanded) + 1):(4 * nrow(i1.new.expanded)),
    c(1:3, 5)] <- cbind.data.frame(newdata.md$S01CaseId_Original, rep(3,
    nrow(i1.new.expanded)), rep("md", nrow(i1.new.expanded)), pseud3, stringsAsFactors = F)
w3 <- predict(w3.model, cbind(newdata.md, pseud.age3_4 = as.factor(pseud3),
    weight.age2_3 = w2)) + rnorm(nrow(newdata.md), mean = 0, sd = sigma(w3.model))
simulated.outcomes.md[(3 * nrow(i1.new.expanded) + 1):(4 * nrow(i1.new.expanded)),
    4] <- w3

# t=4-5
pseud4 <- rbinom(n = nrow(newdata.md), prob = predict(i4.model, cbind(newdata.md,
    pseud.age3_4 = as.factor(pseud3), weight.age3_4 = w3), type = "response"),
    size = 1)
```

```
simulated.outcomes.md[(4 * nrow(i1.new.expanded) + 1):(5 * nrow(i1.new.expanded)),
    c(1:3, 5)] <- cbind.data.frame(newdata.md$S01CaseId_Original, rep(4,
    nrow(i1.new.expanded)), rep("md", nrow(i1.new.expanded)), pseud4, stringsAsFactors = F)
w4 <- predict(w4.model, cbind(newdata.md, pseud.age4_5 = as.factor(pseud4),
    weight.age3_4 = w3)) + rnorm(nrow(newdata.md), mean = 0, sd = sigma(w4.model))
simulated.outcomes.md[(4 * nrow(i1.new.expanded) + 1):(5 * nrow(i1.new.expanded)),
    4] <- w4

# t=5-6
pseud5 <- rbinom(n = nrow(newdata.md), prob = predict(i5.model, cbind(newdata.md,
    pseud.age4_5 = as.factor(pseud4), weight.age4_5 = w4), type = "response"),
    size = 1)
simulated.outcomes.md[(5 * nrow(i1.new.expanded) + 1):(6 * nrow(i1.new.expanded)),
    c(1:3, 5)] <- cbind.data.frame(newdata.md$S01CaseId_Original, rep(5,
    nrow(i1.new.expanded)), rep("md", nrow(i1.new.expanded)), pseud5, stringsAsFactors = F)
w5 <- predict(w5.model, cbind(newdata.md, pseud.age5_6 = as.factor(pseud5),
    weight.age4_5 = w4)) + rnorm(nrow(newdata.md), mean = 0, sd = sigma(w5.model))
simulated.outcomes.md[(5 * nrow(i1.new.expanded) + 1):(6 * nrow(i1.new.expanded)),
    4] <- w5

# t=6-7
pseud6 <- rbinom(n = nrow(newdata.md), prob = predict(i6.model, cbind(newdata.md,
    pseud.age5_6 = as.factor(pseud5), weight.age5_6 = w5), type = "response"),
    size = 1)
simulated.outcomes.md[(6 * nrow(i1.new.expanded) + 1):(7 * nrow(i1.new.expanded)),
    c(1:3, 5)] <- cbind.data.frame(newdata.md$S01CaseId_Original, rep(6,
    nrow(i1.new.expanded)), rep("md", nrow(i1.new.expanded)), pseud6, stringsAsFactors = F)
w6 <- predict(w6.model, cbind(newdata.md, pseud.age6_7 = as.factor(pseud6),
    weight.age5_6 = w5)) + rnorm(nrow(newdata.md), mean = 0, sd = sigma(w6.model))
simulated.outcomes.md[(6 * nrow(i1.new.expanded) + 1):(7 * nrow(i1.new.expanded)),
    4] <- w6

simulated.outcomes.md <- arrange(simulated.outcomes.md, id, age)

#-----------------------------------------------------------------
# simulate weight and infection for exposure = least deprived ---
#-----------------------------------------------------------------
# set up dataset to save results
simulated.outcomes.ld <- as.data.frame(matrix(nrow = 7 * nrow(i1.new.expanded),
    ncol = 5))
colnames(simulated.outcomes.ld) <- c("id", "age", "exposure", "weight", "infection")
# dataset used for simulation
newdata.ld <- select(i1.new.expanded, S01CaseId_Original, dmg_sex, F508_class,
    yob.centered, imd.quintile)
newdata.ld$imd.quintile = "1"

# t=0-1
pseud0 <- rbinom(n = nrow(newdata.ld), prob = predict(i0.model, newdata.ld,
    type = "response"), size = 1)
simulated.outcomes.ld[1:nrow(i1.new.expanded), c(1:3, 5)] <- cbind.data.frame(newdata.ld$S01Cas
    rep(0, nrow(i1.new.expanded)), rep("ld", nrow(i1.new.expanded)), pseud0,
    stringsAsFactors = F)
w0 <- predict(w0.model, cbind(newdata.ld, pseud.age0_1 = as.factor(pseud0))) +
```

```
    rnorm(nrow(newdata.ld), mean = 0, sd = sigma(w0.model))
simulated.outcomes.ld[1:nrow(i1.new.expanded), 4] <- w0

# t=1-2
pseud1 <- rbinom(n = nrow(newdata.ld), prob = predict(i1.model, cbind(newdata.ld,
    pseud.age0_1 = as.factor(pseud0), weight.age0_1 = w0), type = "response"),
    size = 1)
simulated.outcomes.ld[(nrow(i1.new.expanded) + 1):(2 * nrow(i1.new.expanded)),
    c(1:3, 5)] <- cbind.data.frame(newdata.ld$S01CaseId_Original, rep(1,
    nrow(i1.new.expanded)), rep("ld", nrow(i1.new.expanded)), pseud1, stringsAsFactors = F)
w1 <- predict(w1.model, cbind(newdata.ld, pseud.age1_2 = as.factor(pseud1),
    weight.age0_1 = w0)) + rnorm(nrow(newdata.ld), mean = 0, sd = sigma(w1.model))
simulated.outcomes.ld[(nrow(i1.new.expanded) + 1):(2 * nrow(i1.new.expanded)),
    4] <- w1

# t=2-3
pseud2 <- rbinom(n = nrow(newdata.ld), prob = predict(i2.model, cbind(newdata.ld,
    pseud.age1_2 = as.factor(pseud1), weight.age1_2 = w1), type = "response"),
    size = 1)
simulated.outcomes.ld[(2 * nrow(i1.new.expanded) + 1):(3 * nrow(i1.new.expanded)),
    c(1:3, 5)] <- cbind.data.frame(newdata.ld$S01CaseId_Original, rep(2,
    nrow(i1.new.expanded)), rep("ld", nrow(i1.new.expanded)), pseud2, stringsAsFactors = F)
w2 <- predict(w2.model, cbind(newdata.ld, pseud.age2_3 = as.factor(pseud2),
    weight.age1_2 = w1)) + rnorm(nrow(newdata.ld), mean = 0, sd = sigma(w2.model))
simulated.outcomes.ld[(2 * nrow(i1.new.expanded) + 1):(3 * nrow(i1.new.expanded)),
    4] <- w2

# t=3-4
pseud3 <- rbinom(n = nrow(newdata.ld), prob = predict(i3.model, cbind(newdata.ld,
    pseud.age2_3 = as.factor(pseud2), weight.age2_3 = w2), type = "response"),
    size = 1)
simulated.outcomes.ld[(3 * nrow(i1.new.expanded) + 1):(4 * nrow(i1.new.expanded)),
    c(1:3, 5)] <- cbind.data.frame(newdata.ld$S01CaseId_Original, rep(3,
    nrow(i1.new.expanded)), rep("ld", nrow(i1.new.expanded)), pseud3, stringsAsFactors = F)
w3 <- predict(w3.model, cbind(newdata.ld, pseud.age3_4 = as.factor(pseud3),
    weight.age2_3 = w2)) + rnorm(nrow(newdata.ld), mean = 0, sd = sigma(w3.model))
simulated.outcomes.ld[(3 * nrow(i1.new.expanded) + 1):(4 * nrow(i1.new.expanded)),
    4] <- w3

# t=4-5
pseud4 <- rbinom(n = nrow(newdata.ld), prob = predict(i4.model, cbind(newdata.ld,
    pseud.age3_4 = as.factor(pseud3), weight.age3_4 = w3), type = "response"),
    size = 1)
simulated.outcomes.ld[(4 * nrow(i1.new.expanded) + 1):(5 * nrow(i1.new.expanded)),
    c(1:3, 5)] <- cbind.data.frame(newdata.ld$S01CaseId_Original, rep(4,
    nrow(i1.new.expanded)), rep("ld", nrow(i1.new.expanded)), pseud4, stringsAsFactors = F)
w4 <- predict(w4.model, cbind(newdata.ld, pseud.age4_5 = as.factor(pseud4),
    weight.age3_4 = w3)) + rnorm(nrow(newdata.ld), mean = 0, sd = sigma(w4.model))
simulated.outcomes.ld[(4 * nrow(i1.new.expanded) + 1):(5 * nrow(i1.new.expanded)),
    4] <- w4

# t=5-6
pseud5 <- rbinom(n = nrow(newdata.ld), prob = predict(i5.model, cbind(newdata.ld,
```

```r
    pseud.age4_5 = as.factor(pseud4), weight.age4_5 = w4), type = "response"),
    size = 1)
simulated.outcomes.ld[(5 * nrow(i1.new.expanded) + 1):(6 * nrow(i1.new.expanded)),
    c(1:3, 5)] <- cbind.data.frame(newdata.ld$S01CaseId_Original, rep(5,
    nrow(i1.new.expanded)), rep("ld", nrow(i1.new.expanded)), pseud5, stringsAsFactors = F)
w5 <- predict(w5.model, cbind(newdata.ld, pseud.age5_6 = as.factor(pseud5),
    weight.age4_5 = w4)) + rnorm(nrow(newdata.ld), mean = 0, sd = sigma(w5.model))
simulated.outcomes.ld[(5 * nrow(i1.new.expanded) + 1):(6 * nrow(i1.new.expanded)),
    4] <- w5

# t=6-7
pseud6 <- rbinom(n = nrow(newdata.ld), prob = predict(i6.model, cbind(newdata.ld,
    pseud.age5_6 = as.factor(pseud5), weight.age5_6 = w5), type = "response"),
    size = 1)
simulated.outcomes.ld[(6 * nrow(i1.new.expanded) + 1):(7 * nrow(i1.new.expanded)),
    c(1:3, 5)] <- cbind.data.frame(newdata.ld$S01CaseId_Original, rep(6,
    nrow(i1.new.expanded)), rep("ld", nrow(i1.new.expanded)), pseud6, stringsAsFactors = F)
w6 <- predict(w6.model, cbind(newdata.ld, pseud.age6_7 = as.factor(pseud6),
    weight.age5_6 = w5)) + rnorm(nrow(newdata.ld), mean = 0, sd = sigma(w6.model))
simulated.outcomes.ld[(6 * nrow(i1.new.expanded) + 1):(7 * nrow(i1.new.expanded)),
    4] <- w6

simulated.outcomes.ld <- arrange(simulated.outcomes.ld, id, age)

# combine datasets
simulated.outcomes = rbind(simulated.outcomes.ld, simulated.outcomes.md)

expected <- as.data.frame(matrix(ncol = 3, nrow = 25))
colnames(expected) <- c("md.md", "ld.md", "ld.ld")

# combine baseline characteristics with simulated weight and infection
# trajectories for those where exposure was set to most deprived...
simulated.outcomes.md2 <- left_join(simulated.outcomes.md, select(i1.new.expanded,
    S01CaseId_Original, dmg_sex, F508_class, yob.centered), by = c(id = "S01CaseId_Original"))

# .. and least deprived
simulated.outcomes.ld2 <- left_join(simulated.outcomes.ld, select(i1.new.expanded,
    S01CaseId_Original, dmg_sex, F508_class, yob.centered), by = c(id = "S01CaseId_Original"))
simulated.outcomes.ld2 <- arrange(simulated.outcomes.ld2, id, age)

#------------------------------------------------------------
# Create dataset were individuals are exposed (most deprived) but their
# weight trajectory was randomly drawn from the distribution of the
# unexposed (least deprived)
#------------------------------------------------------------
# add simulated weight trajectory under exposure=least deprived to
# dataset where exposure =most deprived
simulated.outcomes.md2$weight.ld.p <- simulated.outcomes.ld2$weight

# update infection trajectory given the weight trajectory under
# exposure=least deprived but epsure=most deprived
simulated.outcomes.md2$updated.infection.ldw <- NA
simulated.outcomes.md2$updated.infection.ldw[simulated.outcomes.md2$age ==
```

```
    0] <- simulated.outcomes.md2$infection[simulated.outcomes.md2$age ==
    0]

# t=1-2
test <- filter(full_join(newdata.md, select(simulated.outcomes.md2, id, age,
    exposure, weight, infection, updated.infection.ldw, weight.ld.p), by = c(S01CaseId_Original
    age == 0)
pseud1 <- rbinom(n = nrow(test), prob = predict(i1.model, cbind(test, pseud.age0_1 = as.factor(
    weight.age0_1 = test$weight.ld.p), type = "response"), size = 1)
simulated.outcomes.md2$updated.infection.ldw[simulated.outcomes.md2$age ==
    1] <- pseud1

# t=2-3
test <- filter(full_join(newdata.md, select(simulated.outcomes.md2, id, age,
    exposure, weight, infection, updated.infection.ldw, weight.ld.p), by = c(S01CaseId_Original
    age == 1)
pseud2 <- rbinom(n = nrow(test), prob = predict(i2.model, cbind(test, pseud.age1_2 = as.factor(
    weight.age1_2 = test$weight.ld.p), type = "response"), size = 1)
simulated.outcomes.md2$updated.infection.ldw[simulated.outcomes.md2$age ==
    2] <- pseud2

# t=3-4
test <- filter(full_join(newdata.md, select(simulated.outcomes.md2, id, age,
    exposure, weight, infection, updated.infection.ldw, weight.ld.p), by = c(S01CaseId_Original
    age == 2)
pseud3 <- rbinom(n = nrow(newdata.md), prob = predict(i3.model, cbind(test,
    pseud.age2_3 = as.factor(test$updated.infection.ldw), weight.age2_3 = test$weight.ld.p),
    type = "response"), size = 1)
simulated.outcomes.md2$updated.infection.ldw[simulated.outcomes.md2$age ==
    3] <- pseud3

# t=4-5
test <- filter(full_join(newdata.md, select(simulated.outcomes.md2, id, age,
    exposure, weight, infection, updated.infection.ldw, weight.ld.p), by = c(S01CaseId_Original
    age == 3)
pseud4 <- rbinom(n = nrow(newdata.md), prob = predict(i4.model, cbind(test,
    pseud.age3_4 = as.factor(test$updated.infection.ldw), weight.age3_4 = test$weight.ld.p),
    type = "response"), size = 1)
simulated.outcomes.md2$updated.infection.ldw[simulated.outcomes.md2$age ==
    4] <- pseud4

# t=5-6
test <- filter(full_join(newdata.md, select(simulated.outcomes.md2, id, age,
    exposure, weight, infection, updated.infection.ldw, weight.ld.p), by = c(S01CaseId_Original
    age == 4)
pseud5 <- rbinom(n = nrow(newdata.md), prob = predict(i5.model, cbind(test,
    pseud.age4_5 = as.factor(test$updated.infection.ldw), weight.age4_5 = test$weight.ld.p),
    type = "response"), size = 1)
simulated.outcomes.md2$updated.infection.ldw[simulated.outcomes.md2$age ==
    5] <- pseud5

# t=6-7
test <- filter(full_join(newdata.md, select(simulated.outcomes.md2, id, age,
```

```r
    exposure, weight, infection, updated.infection.ldw, weight.ld.p), by = c(S01CaseId_Original
    age == 5)
pseud6 <- rbinom(n = nrow(newdata.md), prob = predict(i6.model, cbind(test,
    pseud.age5_6 = as.factor(test$updated.infection.ldw), weight.age5_6 = test$weight.ld.p),
    type = "response"), size = 1)
simulated.outcomes.md2$updated.infection.ldw[simulated.outcomes.md2$age ==
    6] <- pseud6

#----------------------------------------------------------
# simulate outcomes --------------------------------------
#----------------------------------------------------------
# simulated outcomes where exposure=most deprived and weight
# trajectories are randomly drwn from the distribution amongst those
# where exposure=most deprived create wide dataset again to be able to
# simulate from the outcome model
newdata.lung.md.md.1 <- simulated.outcomes.md2 %>%
    group_by(id, age) %>%
    nest(weight, infection, .key = "value_col") %>%
    spread(key = age, value = value_col) %>%
    unnest()
newdata.lung.md.md <- left_join(newdata.lung.md.md.1, unique(select(simulated.outcomes.md2,
    id, dmg_sex, F508_class, yob.centered, exposure)), by = "id")
colnames(newdata.lung.md.md) <- c("id", "weight.age0_1", "pseud.age0_1",
    "weight.age1_2", "pseud.age1_2", "weight.age2_3", "pseud.age2_3", "weight.age3_4",
    "pseud.age3_4", "weight.age4_5", "pseud.age4_5", "weight.age5_6", "pseud.age5_6",
    "weight.age6_7", "pseud.age6_7", "dmg_sex", "F508_class", "yob.centered",
    "imd.quintile")
newdata.lung.md.md$imd.quintile <- "5"

newdata.lung.md.md$pseud.age0_1 <- as.factor(as.character(newdata.lung.md.md$pseud.age0_1))
newdata.lung.md.md$pseud.age1_2 <- as.factor(as.character(newdata.lung.md.md$pseud.age1_2))
newdata.lung.md.md$pseud.age2_3 <- as.factor(as.character(newdata.lung.md.md$pseud.age2_3))
newdata.lung.md.md$pseud.age3_4 <- as.factor(as.character(newdata.lung.md.md$pseud.age3_4))
newdata.lung.md.md$pseud.age4_5 <- as.factor(as.character(newdata.lung.md.md$pseud.age4_5))
newdata.lung.md.md$pseud.age5_6 <- as.factor(as.character(newdata.lung.md.md$pseud.age5_6))
newdata.lung.md.md$pseud.age6_7 <- as.factor(as.character(newdata.lung.md.md$pseud.age6_7))

# simulate lung function
newdata.lung.md.md$lf <- predict(lf.model2, newdata = newdata.lung.md.md) +
    rnorm(nrow(newdata.lung.md.md), mean = 0, sd = sigma(lf.model2))

#----------------------------------------------------
# simulated outcomes where exposure=most deprived but weight
# trajectories are randomly drwn from the distribution amongst those
# where exposure=least deprived create wide dataset again to be able to
# simulate from the outcome model
newdata.lung.md.ld.1 <- simulated.outcomes.md2 %>%
    group_by(id, age) %>%
    nest(weight.ld.p, updated.infection.ldw, .key = "value_col") %>%
    spread(key = age, value = value_col) %>%
    unnest()
newdata.lung.md.ld <- left_join(newdata.lung.md.ld.1, unique(select(simulated.outcomes.md2,
    id, dmg_sex, F508_class, yob.centered, exposure)), by = "id")
```

```r
colnames(newdata.lung.md.ld) <- c("id", "weight.age0_1", "pseud.age0_1",
    "weight.age1_2", "pseud.age1_2", "weight.age2_3", "pseud.age2_3", "weight.age3_4",
    "pseud.age3_4", "weight.age4_5", "pseud.age4_5", "weight.age5_6", "pseud.age5_6",
    "weight.age6_7", "pseud.age6_7", "dmg_sex", "F508_class", "yob.centered",
    "imd.quintile")
newdata.lung.md.ld$imd.quintile <- "5"

newdata.lung.md.ld$pseud.age0_1 <- as.factor(as.character(newdata.lung.md.ld$pseud.age0_1))
newdata.lung.md.ld$pseud.age1_2 <- as.factor(as.character(newdata.lung.md.ld$pseud.age1_2))
newdata.lung.md.ld$pseud.age2_3 <- as.factor(as.character(newdata.lung.md.ld$pseud.age2_3))
newdata.lung.md.ld$pseud.age3_4 <- as.factor(as.character(newdata.lung.md.ld$pseud.age3_4))
newdata.lung.md.ld$pseud.age4_5 <- as.factor(as.character(newdata.lung.md.ld$pseud.age4_5))
newdata.lung.md.ld$pseud.age5_6 <- as.factor(as.character(newdata.lung.md.ld$pseud.age5_6))
newdata.lung.md.ld$pseud.age6_7 <- as.factor(as.character(newdata.lung.md.ld$pseud.age6_7))

# simulate lung function
newdata.lung.md.ld$lf <- predict(lf.model2, newdata = newdata.lung.md.ld) +
    rnorm(nrow(newdata.lung.md.ld), mean = 0, sd = sigma(lf.model2))

#---------------------------------------------------
# simulated outcomes where exposure=least deprived and weight
# trajectories are randomly drwn from the distribution amongst those
# where exposure=least deprived create wide dataset again to be able to
# simulate from the outcome model
newdata.lung.ld.ld.1 <- simulated.outcomes.ld2 %>%
    group_by(id, age) %>%
    nest(weight, infection, .key = "value_col") %>%
    spread(key = age, value = value_col) %>%
    unnest()
newdata.lung.ld.ld <- left_join(newdata.lung.ld.ld.1, unique(select(simulated.outcomes.ld2,
    id, dmg_sex, F508_class, yob.centered, exposure)), by = "id")
colnames(newdata.lung.ld.ld) <- c("id", "weight.age0_1", "pseud.age0_1",
    "weight.age1_2", "pseud.age1_2", "weight.age2_3", "pseud.age2_3", "weight.age3_4",
    "pseud.age3_4", "weight.age4_5", "pseud.age4_5", "weight.age5_6", "pseud.age5_6",
    "weight.age6_7", "pseud.age6_7", "dmg_sex", "F508_class", "yob.centered",
    "imd.quintile")
newdata.lung.ld.ld$imd.quintile <- "1"

newdata.lung.ld.ld$pseud.age0_1 <- as.factor(as.character(newdata.lung.ld.ld$pseud.age0_1))
newdata.lung.ld.ld$pseud.age1_2 <- as.factor(as.character(newdata.lung.ld.ld$pseud.age1_2))
newdata.lung.ld.ld$pseud.age2_3 <- as.factor(as.character(newdata.lung.ld.ld$pseud.age2_3))
newdata.lung.ld.ld$pseud.age3_4 <- as.factor(as.character(newdata.lung.ld.ld$pseud.age3_4))
newdata.lung.ld.ld$pseud.age4_5 <- as.factor(as.character(newdata.lung.ld.ld$pseud.age4_5))
newdata.lung.ld.ld$pseud.age5_6 <- as.factor(as.character(newdata.lung.ld.ld$pseud.age5_6))
newdata.lung.ld.ld$pseud.age6_7 <- as.factor(as.character(newdata.lung.ld.ld$pseud.age6_7))

# simulate lung function
newdata.lung.ld.ld$lf <- predict(lf.model2, newdata = newdata.lung.ld.ld) +
    rnorm(nrow(newdata.lung.ld.ld), mean = 0, sd = sigma(lf.model2))

#-------------------------------------------------
# calculate the total adjusted association and the interventional
# disparity measure direct and indirect effects
```

```r
        #-----------------------------------------------

        # mean outcome in the most deprived if their weight is drawn from the
        # distribution of weight trajectories in the most deprived
        exp.lf.md.md <- mean(newdata.lung.md.md$lf)
        # mean outcome in the most deprived if their weight is drawn from the
        # distribution of weight trajectories in the least deprived
        exp.lf.md.ld <- mean(newdata.lung.md.ld$lf)
        # mean outcome in the least deprived if their weight is drawn from the
        # distribution of weight trajectories in the least deprived
        exp.lf.ld.ld <- mean(newdata.lung.ld.ld$lf)

        # total adjusted association
        te <- exp.lf.ld.ld - exp.lf.md.md
        # interventional disparity measure indirect effect
        idm_ide <- exp.lf.md.ld - exp.lf.md.md
        # interventional disparity measure direct effect
        idm_de <- exp.lf.ld.ld - exp.lf.md.ld

        # save results
        results.boot[z, ] <- c(te, idm_ide, idm_de, exp.lf.md.md, exp.lf.md.ld, exp.lf.ld.ld)
    }
    file.name <- paste("./Results_", imp, ".RData", sep = "")
    save(results.boot, file = file.name)
}
```
